# Coagulation factor XII, XI, and VIII activity levels and secondary events after first ischemic stroke

**DOI:** 10.1101/2019.12.26.19015818

**Authors:** Jessica L. Rohmann, Shufan Huo, Pia S. Sperber, Sophie K. Piper, Frits R. Rosendaal, Peter U. Heuschmann, Matthias Endres, Thomas G. Liman, Bob Siegerink

## Abstract

**Background and Purpose:** Though risk for recurrent vascular events is high following ischemic stroke, little is known about risk factors for secondary events post-stroke. The coagulation factors XII, XI, and VII (FXII, FXI, and FVIII) have already been implicated in first thrombotic events, and our aim was to estimate their effects on vascular outcomes within 3 years after first stroke.

**Methods:** In the PROSpective Cohort with Incident Stroke Berlin (PROSCIS-B) study, we followed participants aged 18 and older for three years after first mild to moderate ischemic stroke event or until occurrence of recurrent stroke, myocardial infarction or all-cause mortality (combined endpoint). High coagulation factor activity levels were compared to normal and low levels, and activities were also analyzed as continuous variables. We used Cox proportional hazards models adjusted for age, sex, and cardiovascular risk factors to estimate hazard ratios (HRs) for the combined endpoint.

**Results:** In total, 92 events occurred in 570 included participants, resulting in an absolute rate of 6.6 events per 100 person-years. After confounding adjustment, high FVIII activity showed the strongest relationship with the combined endpoint (HR=2.05, 95%CI 1.28-3.29). High FXI activity was also associated with an increased risk (HR=1.80, 95%CI 1.09-2.98). Contrarily, high FXII activity was not associated with the combined endpoint (HR=0.86, 95%CI 0.49-1.51). Continuous analyses per standard deviation of each biomarker yielded similar results.

**Conclusions:** In our study of mild to moderate ischemic stroke patients, high activity levels of FXI and FVIII but not FXII were associated with worse vascular outcomes in the three-year period after first ischemic stroke. This is of special interest in light of the ongoing trials of antithrombotic treatments targeting FXI.

## Introduction

Globally, stroke remains a leading cause of disability and mortality.^1,2^ Following first-ever ischemic stroke, risk of secondary events is high.^3–5^ Although many risk factors for first ischemic stroke have been identified, comparatively little is known about factors that contribute to secondary post-stroke events. In fact, a recent systematic review of biomarkers of hemostasis found no conclusive evidence of a single marker ready for use in practice, largely due to the limited number of existing studies.^6^

The coagulation factors XII, XI, and VIII (FXII, FXI, and FVIII) are promising candidates for further investigation in this context. As the initiating factor in the contact activation system, FXII was quickly implicated in first-ever vascular events in animal studies; however, conflicting evidence exists regarding the potential role of FXII in IS events in humans.^7–10^ The role of FXI in hypercoagulability has been more consistently demonstrated, and high levels of FXI have been linked to thrombotic events, especially ischemic stroke.^11–13^ Since FVIII activates thrombin and promotes thrombus formation, it is unsurprising that high levels of FVIII have also been implicated in vascular events.^14,15^ FVIII elevation is observed during the acute phase of stroke as part of the inflammatory response,^16^ however, a dose-dependent relationship between FVIII and thrombosis, independent of this acute phase response, has also been described.^14,15,17,18^ A recent study found that acute ischemic stroke patients with elevated FVIII experienced a higher frequency of recurrent thrombotic events while in-hospital.^19^ However, it remains unknown whether this increased risk due to FVIII elevation also persists in the longer-term for future incident thrombotic events.

In the present study, we aimed to estimate the effects of FXII, FXI, and FVIII activity levels on risk for secondary vascular events among ischemic stroke patients.

## Materials and Methods

### Study population

We used data from the Prospective Cohort with Incident Stroke Berlin (PROSCIS-B; clinicaltrials.gov registration number: NCT01363856). This longitudinal hospital-based observational cohort study has been described in detail elsewhere.^20^ Participants (or legal representatives) provided written informed consent for study participation. The study protocol was approved by the internal review board of the Charité-Universitätsmedizin Berlin (EA1/218/09) and was conducted in accordance with ethical principles described in the Declaration of Helsinki.

In brief, starting in January, 2010, patients aged 18 or older presenting at one of the three tertiary stroke units at the Charité-Universitätsmedizin in Berlin with first-ever stroke defined by WHO criteria^21^ including ischemic stroke, primary hemorrhage, or sinus venous thrombosis, were recruited until February 2013. Participants underwent a baseline visit within one week of the initial event, including a detailed interview, clinical examination and the collection of blood samples stored for later analysis. Participants were contacted annually over a period of three years via telephone interview to document vital status, any incident cardiovascular events and to assess functional outcome. Participants who were not reachable by phone were mailed surveys.

As shown in Fig. 1, for the present study, we excluded non-ischemic stroke patients and patients with severe strokes (defined as National Institute of Health Stroke Scale (NIHSS) of >15). In total, activity measurements for at least one coagulation factor were available for 576 PROSCIS-B participants and subsequently included in these analyses.

**Figure 1.**
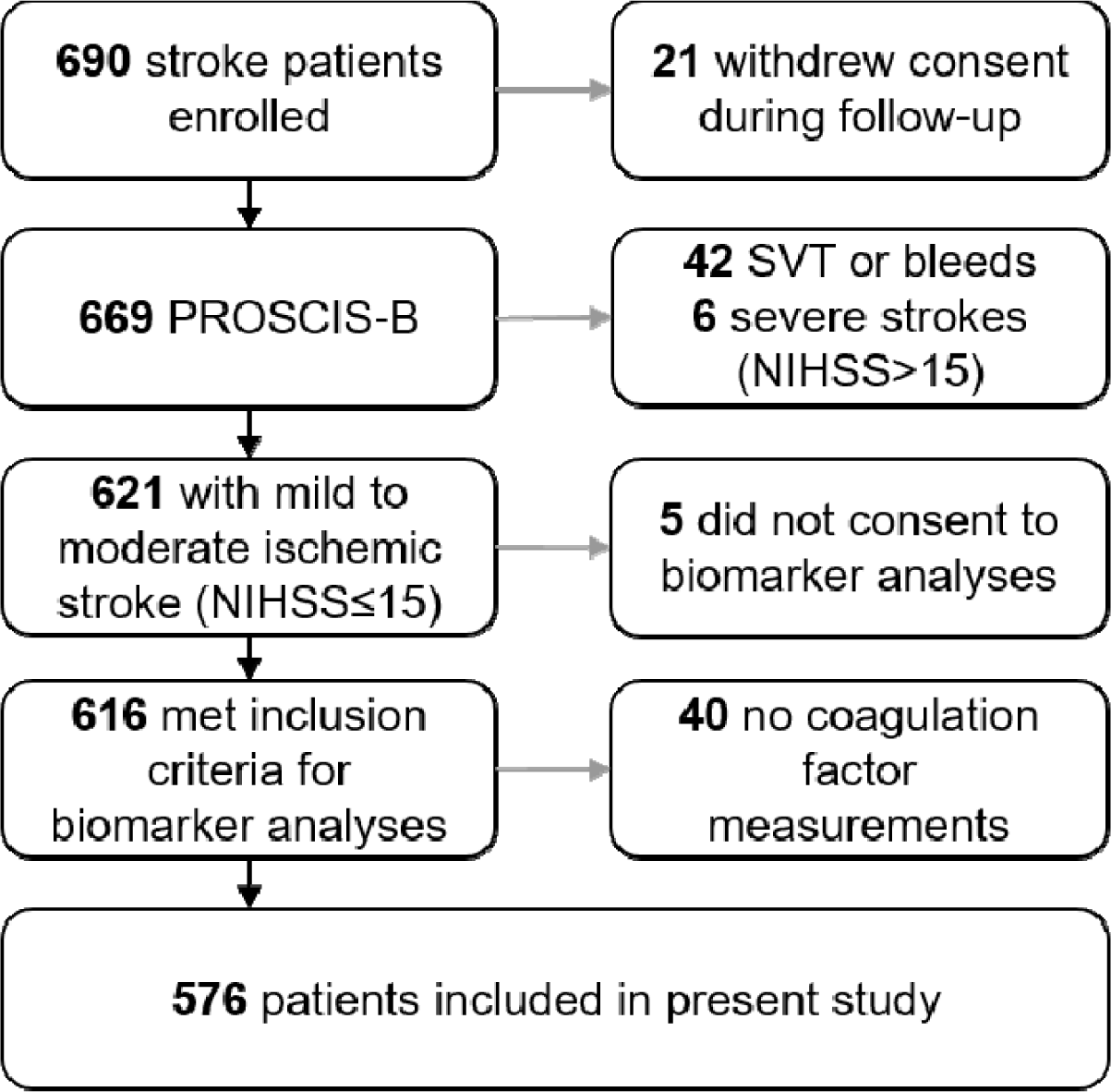
Participant inclusion flowchart. Abbreviations: SVT, sinus venous thrombosis; NIHSS, National Institutes of Health Stroke Scale. Of the 40 participants with no recorded coagulation factor activity measurements, 15 failed in the laboratory and the remainder had no stored citrate available to analyze.

### Participant characteristics

At baseline, age, sex, and cardiovascular risk factors were assessed. In the baseline clinical assessment, body mass index (BMI, in kg/m2), high density lipoprotein (HDL, in mg/dL) and low-density lipoprotein (LDL, in mg/dL) cholesterol were measured. Participants were requested to provide information about lifestyle-related risk factors including smoking (never, former, or current), whether they consumed alcohol regularly, and whether they had a history of diabetes mellitus, hypertension, acute coronary syndrome (myocardial infarction or angina pectoris). The stroke units provided information on whether the patient received thrombolysis, the suspected stroke etiology according to the Trial of ORG 10172 in Acute Stroke Treatment (TOAST) classification^22^, and the severity of the stroke based on the NIHSS (mild: 0-4, moderate 5-15, severe >15).^23^

### Exposure assessment: Coagulation factors

Citrate-buffered blood samples were obtained from PROSCIS-B participants after an overnight fast within 1 week of the initial stroke event and aliquots were stored at −80°C degrees until thawed once for the laboratory assays. Between the initial stroke event and the time of blood sampling, a median of 4 days elapsed (IQR limits: 3-5). Coagulation factor activity levels (:C) were measured using a one-stage clotting assay and are reported as percentages of activated normal pooled plasma (standard activity units). Some of the samples had too little plasma; in these cases, FXI:C followed by FXII:C measurements were prioritized since less is known about these factors in the context of secondary vascular risk compared to FVIII:C. Coagulation factor measurements were performed blinded to participant characteristics or outcome status.

### Outcome: combined endpoint

A combined endpoint outcome was used as the composite of relevant secondary event occurrence; first of either recurrent stroke, myocardial infarction, or death attributable to any cause during follow-up. During follow-up, participants were requested to provide information about the occurrence of any of these events since the last time of contact. These self-reported outcomes were confirmed using the Charité -Universitätsmedizin hospital discharge records or, when not available, information obtained from the treating hospital or general practitioner.

Additional screening of the Charité hospital records was performed to identify any events of interest not self-reported by participants during follow-up. Information about death from any cause was supplemented using city registration office’s records. For one participant, the exact date of death could not be determined and was assigned as the halfway point between last contact and the date on the returned postal questionnaire.

### Endpoint committee

All CV endpoints were confirmed by medical records from the treating hospital or physician and validated by an independent endpoint committee consisting of two senior vascular neurologists. Only these committee-confirmed endpoints were used in our analyses.

### Statistical analysis

For the primary analysis, we categorized the FXI:C, FXII:C and FVIII:C and FXII levels into quartile groups and compared the highest fourth (>75th percentile) to the remainder (reference). In an additional analysis, we analyzed the activity measurements as continuous exposure variables divided by the standard deviation of all measures made for each factor to allow for better comparisons of the estimated effect sizes between the different factors.

For the time-to-event analyses, we calculated person-time from the date of the initial ischemic stroke to the date of occurrence of the combined endpoint (recurrent stroke, myocardial infarction, or death by any cause), loss-to-follow-up, or the study end, whichever came first. We used Kaplan-Meier curves to estimate event-free survivorship and the log-rank test to test for overall crude differences in survivorship curves between groups after visual inspection of fulfillment of the proportional hazards assumption. Dropouts were censored at date of last contact.

We used Cox proportional hazards models to estimate the hazard ratios (HR) and 95% confidence intervals (CI) for the combined endpoint outcome adjusted for potential confounding factors. We used multiple models for confounding control: Model 1 was adjusted only for age and sex. In Model 2, we additionally adjusted for cardiovascular factors determined to contribute to confounding based on *a priori* knowledge. In addition to age and sex, the second model included the continuous variables: BMI, HDL and LDL cholesterol levels; the categorical variable smoking status (never, ever, current), and the following dichotomous variables: regular alcohol consumption, hypertension, diabetes mellitus, and acute coronary syndrome.

As a sensitivity analysis, in a third model, we adjusted for all Model 2 covariates as well as thrombolysis treatment and NIHSS. Since these are both consequences of the stroke, they could be intermediates in the causal path of interest and may not contribute to confounding directly. However, as they may be proxies for pre-stroke confounders, we decided to explore how the effect estimates change with their inclusion in the third model.

We performed all analyses using STATA IC version 14.2 (Stata Corp., College Station, TX, USA).

### Data availability

The original data are available upon reasonable request from the PROSCIS-B principal investigator (TL). The analysis syntax is available upon request from the corresponding author.

## Results

Participant characteristics at baseline are displayed in Table 1. The study population (N=621) was predominantly male (61%), the median age was 69 years (IQR limits: 58-76), and arterial hypertension was observed in 65% and diabetes mellitus in 22% of participants. Twenty percent of participants received thrombolysis therapy. Median activity levels for FXII:C, FXI:C and FVIII:C were 108 (IQR limits: 91-127), 113 (99-130) and 140 (115-166), respectively. At least one of three coagulation factor measurements were available for 576 participants (93%), who were included in the present analyses. All three factor activity measurements were available for 553 participants. All other variables relevant for this study measured at baseline had less than 5% missing values, other than LDL and HDL cholesterol levels, for which 40 participants had missing values.

**Table 1.** Baseline characteristics of PROSCIS-B participants with mild to moderate ischemic stroke.

| PROSCIS-B participants with mild to moderate ischemic stroke (N=621)* |  |  |
| --- | --- | --- |
| Age in years, median (IQR) | 69 | (58-76) |
| Female sex, N (%) | 242 | (39%) |
| BMI in kg/m<sup>2</sup>, median (IQR) | 27 | (24-30) |
| HDL cholesterol in mg/dl, median (IQR) | 49 | (40-60) |
| LDL cholesterol in mg/dl, median (IQR) | 117 | (96-147) |
| Hypertension, N (%) | 406 | (65%) |
| Acute coronary syndrome, N (%) | 99 | (16%) |
| Diabetes mellitus, N (%) | 137 | (22%) |
| Habitual alcohol consumption, N (%) | 217 | (35%) |
| Smoking, N (%) |  |  |
| Former | 201 | (33%) |
| Current | 171 | (28%) |
| TOAST subtype, N (%) |  |  |
| Large-artery atherosclerosis | 167 | (27%) |
| Cardioembolism | 145 | (23%) |
| Small vessel occlusion | 96 | (15%) |
| Other etiology | 22 | (4%) |
| Unknown etiology | 191 | (31%) |
| NIHSS, N (%) |  |  |
| Mild, 0-4 | 470 | (76%) |
| Moderate, 5-15 | 151 | (24%) |
| Lysis therapy, N (%) | 125 | (20%) |
| Coagulation measurements available†, N (%) | 576 | (93%) |
| FXII:C‡, median (IQR) | 108 | (91-127) |
| FXI:C, median (IQR) | 113 | (99-130) |
| FVIII:C, median (IQR) | 140 | (115-166) |

Abbreviations: IQRL, interquartile range limits; BMI, body mass index; HDL, high-density lipoprotein; LDL, low-density lipoprotein; TOAST, stroke etiology according to Trial of Org 10172 in Acute Stroke Treatment; NIHSS, National Institutes of Health Stroke Scale; FVIII:C, coagulation factor VIII activity; FXI:C, coagulation factor XI activity; FXII:C, coagulation factor XII activity.
\*Owing to missing data, percentages may not total 100. All variables have <5% missing values except for the coagulation factor measurements. In total, 34 study participants were missing LDL and HDL cholesterol measurements and 40 participants were missing all three coagulation factor activity measurements.
†Laboratory measurements available for at least one of the coagulation factors of interest. 553 had all three coagulation factor activity measurements.
‡Activities were measured as percentages of activated normal pooled plasma.

After a pursuit follow-up time of 3.0 years resulting in 1,419.5 contributed person-years, 94 combined endpoint events occurred. Of these, 41 were recurrent ischemic strokes, 5 were myocardial infarctions and 48 were deaths. The overall crude observed incidence rate for included participants was 6.6 events per 100 person-years. The absolute cumulative risk for the combined outcome during follow-up among the 576 included participants was 16.3%.

We generated Kaplan-Meier curves to compare participants with coagulation factor levels in the highest fourth (>p75) with the remainder for the three factors of interest (Fig. 2). In the crude comparison, no significant difference between >p75 and ≤p75 groups of FXII:C was observed in the log-rank test (p=0.48). However, clear differences were observed for both the FXI:C and FVIII:C comparisons; participants with high levels had consistently higher cumulative probabilities of the combined endpoint compared to the reference group (FXI:C: p=0.062; FVIII:C: p=0.0001).

**Figure 2.**
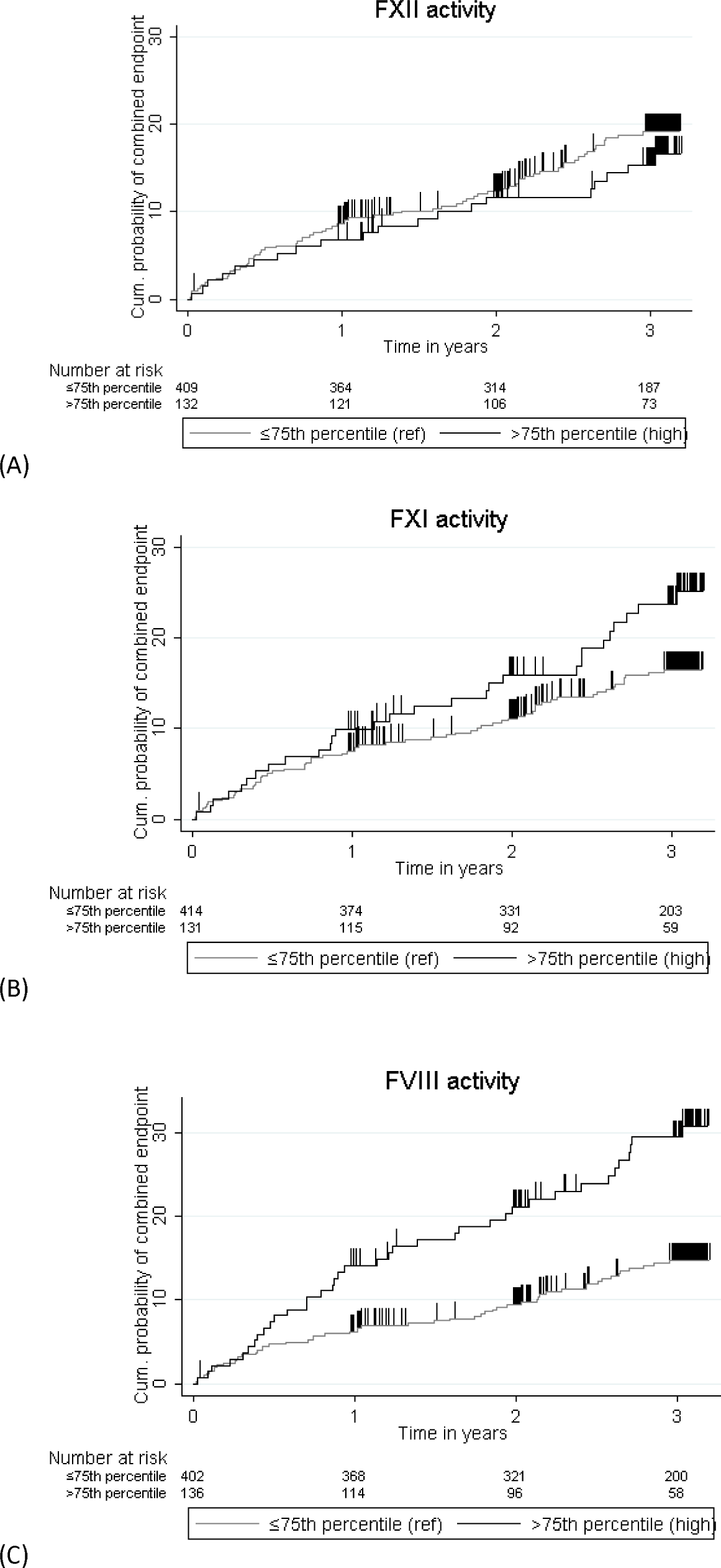
Cumulative probabilities of the combined vascular endpoint. Kaplan-Meier estimates among stroke patient participants with high activity levels (>75th percentile) of each indicated coagulation factor (a) FXII, (b) FXI and (c) FVIII compared to those with lower activity levels of each factor (≤ 75th percentile, reference). Activity levels were measured as percentages of activated normal pooled plasma. Abbreviations: FXI, coagulation factor XI; FXII, coagulation factor XII; FVIII, coagulation factor VIII; cum., cumulative; ref, reference.

The multivariable adjusted HRs are shown in Table 2. In the fully adjusted model (Model 2), high FXII:C levels (>p75) were not associated with the combined endpoint (HR=0.86, 95%CI 0.49-1.51). High FXI:C levels were associated with increased HR for the combined endpoint: (HR=1.80, 95%CI 1.09-2.98), as were high FVIII:C levels (HR=2.05, 95%CI 1.28-3.29). In the secondary analyses treating the coagulation factor levels as continuous variable, similar results were obtained (Table 2). One standard deviation of FXII:C, FXI:C and FVIII:C levels corresponded to 29.3, 28.8, and 45.4 units, respectively.

**Table 2.**
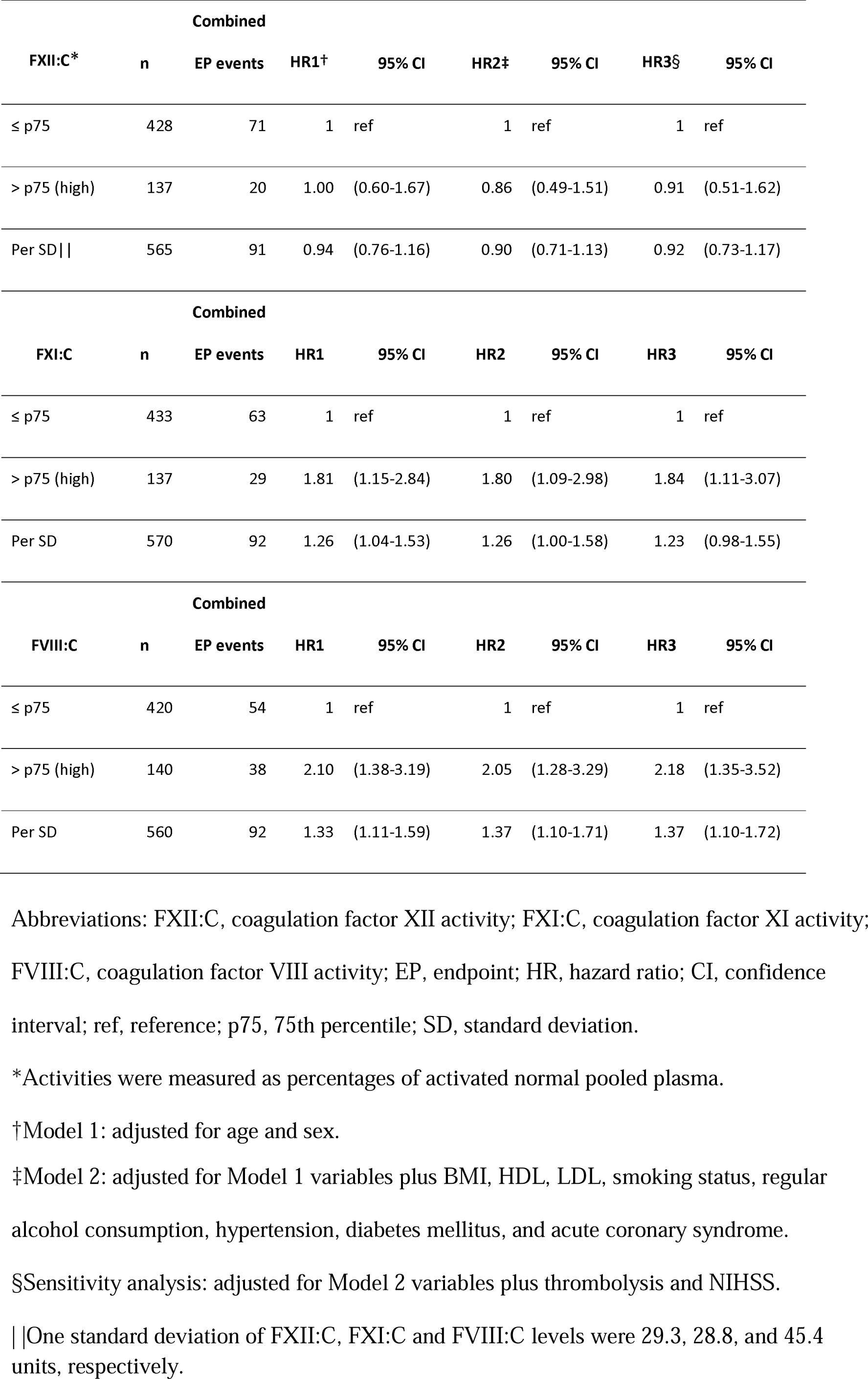
Hazard ratios from Cox proportional hazards regression models for FXII:C, FXI:C.

In a sensitivity analysis, we further adjusted Model 2 for NIHSS and thrombolysis. This additional adjustment did not substantially change the results (Table 2).

## Discussion

In this prospective patient cohort study of individuals with mild to moderate ischemic stroke, having high levels of FXI or FVIII activity was associated with an increased hazard for the combined vascular endpoint within three years compared to individuals with low/normal levels after confounding adjustment. High FXII activity did appear to be associated with a strong increase or decrease in hazard for the combined endpoint.

Though the relationships between coagulation factors FXII, FXI and FVIII and primary thrombotic event risk, for both venous and arterial events, have been well-studied, to the best of our knowledge, this is the first study to explore their role in long-term vascular event risk after first stroke. Furthermore, a limited number of studies have investigated the impact of multiple coagulation factors in a single study. Our findings expand on recent literature findings and fill some important gaps with longitudinal results.

Specifically in the context of post-stroke outcomes, the search for meaningful etiologic or prognostic hemostatic biomarkers has not been straightforward. A recent systematic review of hemostatic biomarkers in ischemic stroke revealed a large heterogeneity in existing studies and did not find enough evidence to provide clear recommendations for a prognostic marker to be used in practice, however, some biomarkers, including FXI and FVIII, seemed promising in some of the included studies.^6^

Data from our study is in line with findings from a cross-sectional study, which concluded that active FXI in plasma may be linked to worse outcomes in patients with acute cerebrovascular events.^24^

Another study found that acute ischemic stroke patients with elevated FVIII experienced a higher frequency of recurrent thrombotic events while in-hospital^19^ and our findings add that this relationship persists in the longer-term for future incident vascular events. Regarding ischemic stroke patients who underwent thrombolysis, a study found that having elevated FVIII levels, both immediately and 24 hours after thrombolysis, led to a higher risk for poor functional outcome (mRS□≥□3) at 90□days.^25^

There have been some important recent developments regarding hemostatic factors in the context of first stroke that may also inform future treatment strategies in the context of secondary prevention. For instance, FXI appears to be a stronger risk factor for first ischemic stroke than myocardial infarction^26^ and may therefore be a particularly attractive target.

Numerous laboratory and animal studies have further demonstrated the thrombotic role of FXII and FXI in thrombosis but non-integral role of these factors in normal hemostasis, making them promising targets for future anticoagulant drug development with potentially lower bleeding risk.^7,27,28^ In addition to the successful trial conducted among knee arthroplasty patients, in which FXI antisense oligonucleotides prevented venous thrombotic events without increasing bleeding risk,^29^ a recent genetic study investigating variants known to alter FXI levels and increase relative activated partial thromboplastin time found that genetic disposition to lower FXI levels was associated with lower odds for ischemic stroke without increasing risk for major bleeding.^30^ This decrease was equivalent to the FXI level reduction that can be achieved through pharmacological modulation.^30^

Furthermore, a 2018 study using two-sample Mendelian randomization found that genetically determined FXI levels had a causal effect on risk of any ischemic stroke but not myocardial infarction or intracerebral hemorrhage, with the strongest effect observed amongst the cardioembolism subgroup.^31^ Shortly after, a 2019 study also using Mendelian randomization techniques in a larger meta-analysis integrating phenomic, genomic and proteomic databases assessed the role of 653 proteins as potential mediators for ischemic stroke subtypes and potential relevant side effects and linked diseases of hypothetical treatment strategies.^32^ In this study, genetically determined FXI levels were identified as one of five causal mediators of ischemic stroke, with the cardioembolic subtype appearing to drive this effect.^32^ In both studies, no adverse side effects appeared to be linked to the genetic influences on variation in FXI levels, providing further justification for clinical trials on FXI-related interventions in the context of ischemic stroke.^32^

In light of our findings that stroke patients with high FXI:C had a higher risk for secondary events after first stroke, the population of stroke patients with high FXI levels may particularly benefit from such interventions, and FXI:C may be a useful biomarker in identifying individuals who are most likely to benefit from such interventions.

### Strengths and Limitations

Strengths of our study include the large number of participants given the difficult-to-reach nature of the stroke patient population. Coagulation activity was assayed on state of the art machinery on fresh-frozen, once-thawed plasma samples as opposed to antigen level measurements. Furthermore, our longitudinal design with 94 observed outcome events afforded us the opportunity to contribute our analyses of long-term outcome risk to the literature currently limited to cross-sectional and very-short-term designs.

We were able to screen for additional, unreported clinical endpoints of interest in the Charité University Hospital medical records to supplement the information provided by the patients, however, unreported clinical endpoints presenting at other clinics in Berlin or elsewhere may have been missed. When possible, we made an effort to validate any patient-reported events by requesting forwarding of medical records from other hospitals and clinics. We further confirmed the vital status for all participants at the end of the study via the local citizen’s registration office in Berlin; however, due to legal restrictions, specific cause information from death certificates was not obtainable.

Some limitations should also be considered when interpreting our results. First, self-reported patient characteristics, such as the lifestyle-related factors and the presence of chronic diseases at baseline, may be prone to recall bias. A set of standard operating procedures and training was provided for the study nurses in an effort to improve consistency in the variables measured at study enrollment. Although we believe that we have included the most important potential sources of confounding in the adjusted models, we cannot rule out that some residual confounding may be present due to unmeasured factors.

Second, we emphasize that the coagulation factor activity levels were measured in blood samples that were taken after the index stroke event. Though FXII and FXI are not known to change dramatically during the acute phase of stroke, this phenomenon has been well-documented for FVIII. This means that our findings for FVIII are likely a mixing of elevated FVIII as part of the acute phase and high pre-stroke FVIII levels (increase of thrombotic event risk independent of the acute phase^18^). It is also possible that these levels changed within the first week post-stroke during which the blood was drawn. Future confirmatory studies should consider time-standardized measurements; sequential measurements could provide additional insights into the changes that occur shortly after stroke.

Our reported results apply to a cohort comprised of first-ever mild to moderate ischemic stroke patients. The 6 patients enrolled with a baseline NIHSS >15 in PROSCIS-B study were excluded limit heterogeneity of the cohort. Readers should take care not to extrapolate our conclusions to severe patients (NIHSS >15).

We do not expect that the censoring of individuals lost to follow up was differential with respect to exposure status. However, it is feasible despite our efforts to confirm unreported endpoints that those who were lost to follow up may have been more likely to experience one of the combined vascular endpoints compared to the participants actively remaining in the study.

## Conclusions

Our study of mild to moderate ischemic stroke patients indicates that high levels of FXI:C or FVIII:C measured within one week of the index event may contribute to unfavorable vascular outcomes after stroke in the longer term (three years). We did not observe a clear relationship with FXII:C. Further research in this area should focus on obtaining time-standardized and repeated measures of coagulation factor activities after stroke. In the context of secondary prevention, we demonstrated that individuals with high levels of FXI:C after stroke have an increased risk for secondary events. This knowledge may be beneficial for potential future treatment strategies involving drugs targeting FXI.

## Acknowledgements

We thank P. Noordijk, L. Mahic and A. Hoenderdos from the Leiden University Medical Center for performing laboratory assays and J. Thümmler for PROSCIS-B data management. We are indebted to the participants of the PROSCIS-B for their commitment. JLR is a PhD candidate at the Charité – Universitätsmedizin Berlin, and this work is submitted in partial fulfillment of her PhD requirements.

## References

1. Institute for Health Metrics and Evaluation (IHME). Seattle, WA: IHME, University of Washington. GBD Compare Data Visualization [Internet]. 2018 [cited 2019 Feb 28];Available from: http://vizhub.healthdata.org/gbd-compare

2. Thrift AG, Thayabaranathan T, Howard G, Howard VJ, Rothwell PM, Feigin VL, et al. Global stroke statistics. Int. J. Stroke. 2017;12:13–32.

3. Mohan KM, Wolfe CDA, Rudd AG, Heuschmann PU, Kolominsky-Rabas PL, Grieve AP. Risk and cumulative risk of stroke recurrence: a systematic review and meta-analysis. Stroke. 2011;42:1489–1494.

4. Dennis MS, Burn JP, Sandercock PA, Bamford JM, Wade DT, Warlow CP. Long-term survival after first-ever stroke: the Oxfordshire Community Stroke Project. Stroke. 1993;24:796–800.

5. Sacco RL, Wolf PA, Kannel WB, McNamara PM. Survival and recurrence following stroke. The Framingham study. Stroke. 1982;13:290–295.

6. Donkel SJ, Benaddi B, Dippel DWJ, Ten Cate H, de Maat MPM. Prognostic Hemostasis Biomarkers in Acute Ischemic Stroke. Arterioscler. Thromb. Vasc. Biol. 2019;39:360–372.

7. Gailani D, Bane CE, Gruber A. Factor XI and contact activation as targets for antithrombotic therapy. J. Thromb. Haemost. 2015;13:1383–1395.

8. Kraft P, De Meyer SF, Kleinschnitz C. Next-generation antithrombotics in ischemic stroke: preclinical perspective on “bleeding-free antithrombosis.” J. Cereb. Blood Flow Metab. 2012;32:1831–1840.

9. Siegerink B, Govers-Riemslag JWP, Rosendaal FR, Ten Cate H, Algra A. Intrinsic coagulation activation and the risk of arterial thrombosis in young women: results from the Risk of Arterial Thrombosis in relation to Oral contraceptives (RATIO) case-control study. Circulation. 2010;122:1854–1861.

10. Govers-Riemslag JWP, Smid M, Cooper JA, Bauer KA, Rosenberg RD, Hack CE, et al. The plasma kallikrein-kinin system and risk of cardiovascular disease in men. J. Thromb. Haemost. 2007;5:1896–1903.

11. Yang DT, Flanders MM, Kim H, Rodgers GM. Elevated factor XI activity levels are associated with an increased odds ratio for cerebrovascular events. Am. J. Clin. Pathol. 2006;126:411–415.

12. Suri MFK, Yamagishi K, Aleksic N, Hannan PJ, Folsom AR. Novel hemostatic factor levels and risk of ischemic stroke: the Atherosclerosis Risk in Communities (ARIC) Study. Cerebrovasc. Dis. 2010;29:497–502.

13. Siegerink B, Rosendaal FR, Algra A. Antigen levels of coagulation factor XII, coagulation factor XI and prekallikrein, and the risk of myocardial infarction and ischemic stroke in young women. J. Thromb. Haemost. 2014;12:606–613.

14. Borissoff JI, Spronk HMH, ten Cate H. The hemostatic system as a modulator of atherosclerosis. N. Engl. J. Med. 2011;364:1746–1760.

15. Siegler JE, Samai A, Albright KC, Boehme AK, Martin-Schild S. Factoring in Factor VIII With Acute Ischemic Stroke. Clin. Appl. Thromb. Hemost. 2015;21:597–602.

16. Chang TR, Albright KC, Boehme AK, Dorsey A, Sartor EA, Kruse-Jarres R, et al. Factor VIII in the setting of acute ischemic stroke among patients with suspected hypercoagulable state. Clin. Appl. Thromb. Hemost. 2014;20:124–128.

17. Koster T, Blann AD, Briët E, Vandenbroucke JP, Rosendaal FR. Role of clotting factor VIII in effect of von Willebrand factor on occurrence of deep-vein thrombosis. Lancet. 1995;345:152–155.

18. Kamphuisen PW, Eikenboom JC, Bertina RM. Elevated factor VIII levels and the risk of thrombosis. Arterioscler. Thromb. Vasc. Biol. 2001;21:731–738.

19. Gouse BM, Boehme AK, Monlezun DJ, Siegler JE, George AJ, Brag K, et al. New Thrombotic Events in Ischemic Stroke Patients with Elevated Factor VIII. Thrombosis. 2014;2014:302861.

20. Liman TG, Zietemann V, Wiedmann S, Jungehuelsing GJ, Endres M, Wollenweber FA, et al. Prediction of vascular risk after stroke - protocol and pilot data of the Prospective Cohort with Incident Stroke (PROSCIS). Int. J. Stroke. 2013;8:484–490.

21. Hatano S. Experience from a multicentre stroke register: a preliminary report. Bull. World Health Organ. 1976;54:541–553.

22. Adams HP Jr, Bendixen BH, Kappelle LJ, Biller J, Love BB, Gordon DL, et al. Classification of subtype of acute ischemic stroke. Definitions for use in a multicenter clinical trial. TOAST. Trial of Org 10172 in Acute Stroke Treatment. Stroke. 1993;24:35–41.

23. Lyden P. Using the National Institutes of Health Stroke Scale: A Cautionary Tale. Stroke. 2017;48:513–519.

24. Undas A, Slowik A, Gissel M, Mann KG, Butenas S. Active tissue factor and activated factor XI in patients with acute ischemic cerebrovascular events. Eur. J. Clin. Invest. 2012;42:123–129.

25. Tóth NK, Székely EG, Czuriga-Kovács KR, Sarkady F, Nagy O, Lánczi LI, et al. Elevated Factor VIII and von Willebrand Factor Levels Predict Unfavorable Outcome in Stroke Patients Treated with Intravenous Thrombolysis. Front. Neurol. 2017;8:721.

26. Maino A, Rosendaal FR, Algra A, Peyvandi F, Siegerink B. Hypercoagulability Is a Stronger Risk Factor for Ischaemic Stroke than for Myocardial Infarction: A Systematic Review. PLoS One. 2015;10:e0133523.

27. Weitz JI, Fredenburgh JC. Factors XI and XII as Targets for New Anticoagulants [Internet]. Frontiers in Medicine. 2017;4. Available from: http://dx.doi.org/10.3389/fmed.2017.00019

28. van Montfoort ML, Meijers JCM. Anticoagulation beyond direct thrombin and factor Xa inhibitors: indications for targeting the intrinsic pathway?Thromb. Haemost. 2013;110:223–232.

29. Büller HR, Bethune C, Bhanot S, Gailani D, Monia BP, Raskob GE, et al. Factor XI antisense oligonucleotide for prevention of venous thrombosis. N. Engl. J. Med. 2015;372:232–240.

30. Georgi B, Mielke J, Chaffin M, Khera AV, Gelis L, Mundl H, et al. Leveraging Human Genetics to Estimate Clinical Risk Reductions Achievable by Inhibiting Factor XI. Stroke. 2019;50:3004–3012.

31. Gill D, Georgakis MK, Laffan M, Sabater-Lleal M, Malik R, Tzoulaki I, et al. Genetically Determined FXI (Factor XI) Levels and Risk of Stroke. Stroke. 2018;49:2761–2763.

32. Chong M, Sjaarda J, Pigeyre M, Mohammadi-Shemirani P, Lali R, Shoamanesh A, et al. Novel Drug Targets for Ischemic Stroke Identified Through Mendelian Randomization Analysis of the Blood Proteome. Circulation. 2019;140:819–830.

